# Nowcasting and Forecasting COVID-19 Waves: The Recursive and Stochastic Nature of Transmission

**DOI:** 10.1101/2022.04.12.22273804

**Authors:** Vinicius V.L. Albani, Roseane Albani, Eduardo Massad, Jorge P. Zubelli

## Abstract

We propose a parsimonious, yet effective, susceptible-exposed-infected-removed-type model that incorporates the time change in the transmission and death rates. The model is calibrated by Tikhonov-type regularization from official reports from New York City (NYC), Chicago, the State of São Paulo, in Brazil, and British Columbia, in Canada. To forecast, we propose different ways to extend the transmission parameter, considering its estimated values. The forecast accuracy is then evaluated using real data from the above referred places. All the techniques accurately provided forecast scenarios for periods 15 days long. One of the models effectively predicted the magnitude of the four waves of infections in NYC, including the one caused by the Omicron variant for periods of 45 days long using out-of-sample data.

## 1 Introduction

Since the seminal work of Kermack-McKendrick [1], susceptible-infected-removed-type and susceptible-exposed-infected-removed-type (SEIR-type) models have played a crucial role in understanding the dynamics of infectious diseases through nowcasting and helping public health authorities to address disease outbreaks by forecasting [2]. In the pandemic of COVID-19, that started in the end of 2019 and was still ongoing in the beginning of 2022, SEIR-type models were again recurrently used to nowcast the SARS-CoV-2 spread dynamics and to forecasting [3–12].

Two of the main challenges the COVID-19 pandemic posed in the modeling of infectious diseases were to provide accurate nowcast and forecasts [9,13,14]. Nowcasting is fundamental to understanding the seriousness of the present situation based on the observed reports of infections and deaths, as well as the hospital capacity. Forecasting is the main tool used to inform how much intervention will be needed in the future to control or contain the pathogen spread. Thus, to address both tasks, it is necessary to access good quality data and to design accurate models that incorporate the time evolution of the transmission pattern as well as the disease severity. Moreover, it is necessary to use model calibration techniques that incorporate the uncertainties in data acquisition.

The way a pathogen spreads changes with time since it can be affected, for example, by the adoption of spread control measures and changes in the pathogen (e.g. mutation). However, the way such changes in the pathogen occur is in general unknown and hard to predict. In other words, transmission can be defined as a stochastic process. Thus, to design trustworthy and accurate scenarios, a model must incorporate such variability and randomness. Moreover, based on the past information, the model must be able to predict the future pattern of transmission, presenting realistic potential numbers of infections and deaths. A model that fails to address the latter appropriately can, for example, give arguments to make the public opinion skeptical about or even to deny the severity of an emerging disease. This seemed to be an important issue in the beginning of the COVID-19 pandemic when a series of predictions made with over-simplistic models and calibrated with poor data provided unrealistic catastrophic scenarios [13, 15]. During secondary outbreaks, the implementation of non-pharmaceutical contention measures became more difficult and anti-lockdown protests emerged worldwide [16–18].

Due to these issues in model predictions, forecast accuracy was recurrently discussed during the COVID-19 pandemic. For example, Ioannidis et al. [13] recalls a series of unrealistic scenarios designed in the beginning of the pandemic and proposed some ways to improve model forecast. Gnanvi et al. [19] made a systematic review of models used to predict scenarios of infections and deaths related to COVID-19. They classified their results by comparing predictions with reports and found that reports were inside the 95% confidence intervals of 75% of the considered models. Other works, proposed to refine the modeling of the dynamics of COVID-19 by incorporating, for example, the spatial distribution of infections, mobility information, and different levels of disease severity depending on age and sex [3, 10, 20–23].

To address the issues raised above, the present article proposes a methodology to appropriately model, calibrate, and forecast scenarios using SEIR-type models. More precisely, we show that even simpler SEIR-type models with appropriately calibrated time-dependent transmission parameters are still suitable to forecast scenarios accurately. Moreover, we compare the results of a series of techniques used to define the future pattern of transmission based on past information. All the results are based on real data from Chicago, New York City (NYC), the Canadian province of British Columbia, and the Brazilian state of São Paulo. Some of the techniques were able to predict accurately four large outbreaks in NYC, including those caused by the Delta and Omicron variants of the SARS-Cov-2 using out-of-sample data. Another implication of the numerical results is that considering the pathogen transmission as a stochastic process can lead to more accurate scenarios.

## 2 Methods

As pointed in a series of recent works [3, 5, 24, 25], the estimated values of the transmission parameter usually denoted by *β* change with time and presents some sort of randomness. To make accurate predictions, such variability must be incorporated into the model. Thus, the techniques proposed in what follows are designed to accurately predict the future pattern of transmission by accounting for past calibrated values of the time-dependent *β*. It is worth mentioning that using time-dependent transmission parameters also allow a remarkable adherence to reported data [3, 5, 24, 25].

### 2.1 Describing the Disease Spread Dynamics

To describe the spread dynamics of the virus SARS-CoV-2, we consider a SEIR-type model that, for simplicity, does not consider natural deaths and births. The model accounts for the following five compartments, susceptible (S), exposed (E), infected (I), recovered (R), and deceased (D). The progress between the compartments is defined by the dynamical system of ordinary differential equations below [26]:

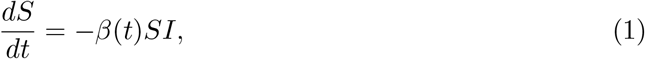

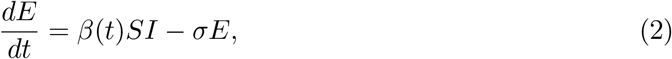

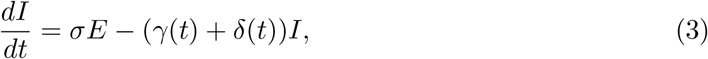

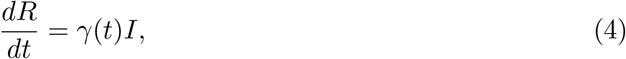

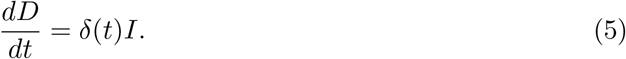

The time-dependent transmission parameter is denoted by *β*(*t*) and the infection to onset mean time is represented by 1*/σ*, which is set to 5.1 days [27]. The quantity *γ* denotes the recovery rate and *δ*(*t*) is the death rate. Following [28], we set *γ* = 1*/*14 days^*−*1^. For each day *t*, the death rate *δ*(*t*) is defined as follows, it is the total number of registered deaths on day *t* divided by the number of reported infection on the day *t −* 12. So it is dimensionless. The 12 days is the meantime that someone takes from being infected to die by COVID-19 [3, 24, 25].

Natural death and birth could be included in the model, but such terms would make little difference in the analysis that follows, as the time interval considered here is small. So, to keep the model as simple as possible, we decided to not include such terms.

### 2.2 Model Calibration

The estimation of the transmission parameter *β* from daily reports of COVID-19 infections is performed by the minimization of the Tikhonov-type functional below for each day *t*, as in [3]:

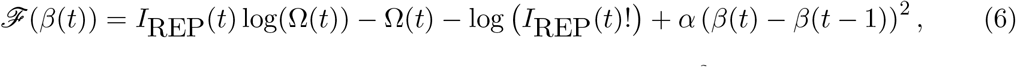

where *α* is the regularization parameter, which has dimension days^2^, to make the second term in the right-hand side of Eq. (6) dimensionless, and *I*REP(*t*) denotes the reported COVID-19 infections on the day *t*. The term log (*I*REP(*t*)!) is approximated by the Stirling’s formula:

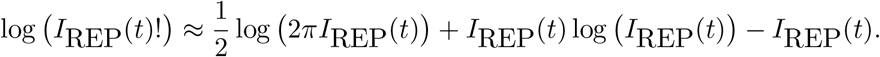

The quantity 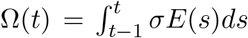 in Eq. (6) represents the model predicted infections for the day *t*. It depends on *β*, since it is part of the solution of the system of equations in Eqs. (2)–(5). Therefore, the *β*(*t*) that minimizes Eq. (6) is the value that makes the model to predict correctly the infections reported in the day *t*. The first part of the functional in Eq. (6) accounts for the discrepancy between the model predictions and the reports of infections. The second part penalizes the first to avoid overfitting, by imposing that the discrepancy between *β*(*t*) and the already known value *β*(*t −* 1) must also be minimized. To avoid the introduction of bias in the calibration process, the parameter *α* must be appropriately chosen. In the present set of examples, we set *α* = 10^*−*3^*/*2 accordingly to an *a posteriori* rule [29].

### 2.3 Model Forecast Techniques

The transmission parameter *β*, as we shall see, changes with time, presenting an uncertain pattern. Since we do not have any formulation to predict how *β* changes with time, we propose five different techniques to evaluate future values of beta allowing the SEIR-type model to make predictions.

#### Average Method (AM)

The simplest technique to extend *β* by setting its future values as the average of its estimated values. More precisely, for any *t > t*_*n*_, set 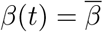, where

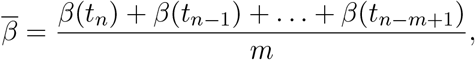

*t*_*n−m*+1_, …, *t*_*n*_ are time values where *β* is calibrated, and *m < n*.

Since the trend of *β* at the end of the dataset is the most relevant information to predict the number of infections in the following days, we set *m* = 5 days. If *m* is large, it is possible to corrupt predictions with outdated information. Moreover, keeping the number of unknowns considerably smaller than the size of “observed” quantities can help to reduce the effect of noise in the predictions.

#### Linear Regression (LR)

Another simple technique to extend *β* is to fit a linear model. In other words, for any *t > t*_*n*_, we set *β*(*t*) = *a · t* + *b*, with the scalar parameters *a* and *b* fitted to the set of calibrated values of *β {β*(*t*_*n−m*+1_), *β*(*t*_*n−m*+2_), …, *β*(*t*_*n*_)*}*. The calibration is performed by least-squares.

In this case, since there are two unknowns, we set *m* = 10 days also to avoid outdated information and to keep the number of unknowns considerably smaller than the number of observations.

#### Truncated Fourier Series (TFS)

A more elaborated technique is to calibrate a truncated Fourier series [30] that approximates a dataset. More precisely, assume that *β* can be approximated by a truncated Fourier series in the time interval [*t*_*n−m*+1_, *t*_*n*_],

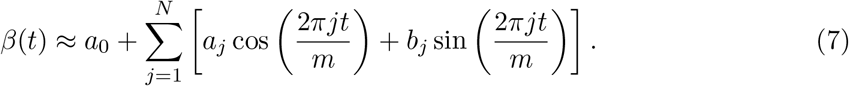

For *m* and *N* fixed, the parameters *a*_0_, *a*_*j*_, and *b*_*j*_ (*j* = 1, …, *N*), which have the same dimension of *β*, are calibrated by least squares from the dataset *{β*(*t*_*n−m*+1_), *β*(*t*_*n−m*+2_), …, *β*(*t*_*n*_)*}*. Thus, for each time *t > t*_*n*_, *β*(*t*) is given by the periodic function defined in Eq. (7) with calibrated parameters.

In this case, choosing the length of the time interval where the series is defined and the number of terms in the series is tricky. After testing different values and configurations, we found that setting the length of the time interval *m* = 30 days and the number of terms in the series *N* = 3 gives a computational affordable technique that fits well the data.

#### Truncated Fourier Series with Noise (TFSWN)

In order to add some uncertainty to the predicted values of *β*, we add to the truncated Fourier series described above a Gaussian noise with mean zero and standard deviation given as follows:

1. Evaluate the difference between the values given by the truncated Fourier series and the dataset *{β*(*t*_*n−m*+1_), *β*(*t*_*n−m*+2_), …, *β*(*t*_*n*_)*}*.
2. Evaluate the empirical standard deviation of the difference.

For each time *t > t*_*n*_, *β*(*t*) is given by the function in Eq. (7) plus a Gaussian random variable with mean zero and standard deviation given as above. In this case, we use the same values for *m* and *N*, as well as the parameters of the truncated Fourier series in Eq. (7) found for the noiseless case.

#### A Mean-Reverting (MR) Model for the Transmission Parameter

As an alternative to the methods described above, we propose a mean-reverting model for the dynamics of *β*. Since the time evolution of *β* seems to return to mean values, after random changes, it is natural to test such kind of model. So, inspired by the volatility of the Heston model [31] from Quantitative Finance, we assume that, for *t > t*_*n*_, the values of *β*(*t*) are given by the following stochastic process

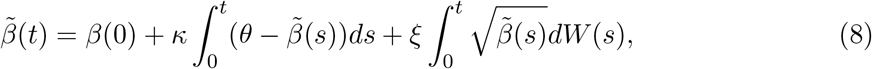

where *{W* (*s*) : *s >* 0*}* represents the Wiener process also known as Brownian motion. Thus, set 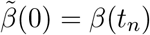, and 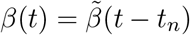.

Based on the Euler-Maruyama scheme to solve the stochastic differential equation in Eq. (8), we have,

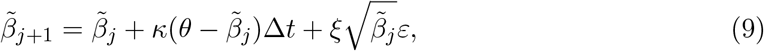

where *ε* has Gaussian distribution with mean zero and variance Δ*t*.

Since the estimated values for *β* are given daily, we set Δ*t* = 1*/*365. The parameters *κ, θ*, and *ξ* are obtained from the set of estimated values for *β, {β*(*t*_*n−m*+1_), *β*(*t*_*n−m*+2_), …, *β*(*t*_*n*_)*}*, by minimizing the functional

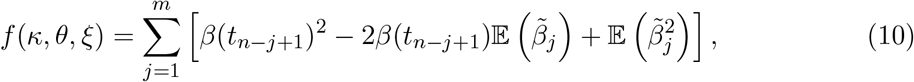

with respect to the parameters *κ, θ, ξ*, where 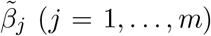 is a function of *κ, θ, ξ*. It is worth noticing that, in the calibration,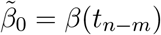.

The quantities 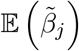 and 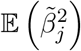 are deterministic and can be easily evaluated by the following recursive formulas:

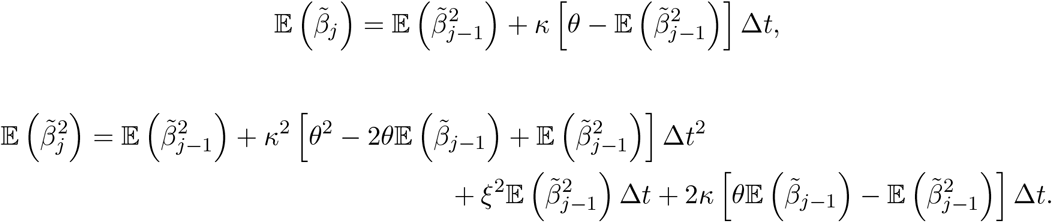

In the results, we shall use *m* = 30 days, which keeps the number of unknowns much smaller than the number of observations, and, in principle, allows the model to incorporate the most recent data.

## 3 Results

Next, we perform a series of numerical tests to compare the accuracy of the predicted accumulated numbers of infections obtained using the techniques described in Section 2.

Firstly, the SEIR-type model in Eqs. (2)–(5) is calibrated using 7-day moving average time-series of the daily reports of COVID-19 infections and deaths from Chicago and New York City(NYC) in the United States (US), the Canadian province of British Columbia (BC), and the Brazilian state of São Paulo (SP). The datasets start mainly in the beginning of 2020 and end at the end of 2021.

We consider predictions of infections for 10, 15, 30, 45, 60, and 90 days after the period used in the calibration. Predictions are made as, follows: we calibrate the model until some date *t*_*n*_ using the reported infections until the date *t*_*n*_, then, for any *t*_*j*_ *> t*_*n*_ we evaluate the model predictions of infections using the five techniques proposed in Section 2. These predictions are updated daily until the end of the dataset. We compare the reported and predicted accumulated numbers of infections, evaluating the normalized error in the out-of-sample predictions as follows, denote by *I*REP the accumulated number of reported infections during the time interval of forecast and by *I*PRED the accumulated predicted infections for the same period, then,

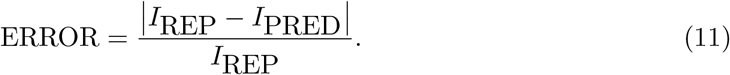

The error is evaluated for the daily out-of-sample predictions and classified by the number of days ahead and the technique used to generate the prediction. by out-of-sample we mean that the model calibration was performed with a dataset different from the one used to compare the model predictions. The daily errors are then used to evaluate median values and 70% confidence intervals (70% CI). The comparison between the errors median values can be found in Figure 1. Table 1 presents the errors median values and their 70% CI.

**Table 1:** Median values and their corresponding 70% confidence intervals of the percentage error in the out-of-sample predicted accumulated numbers of infections. Predictions are based on calibrated data and evaluated from 10 to 90 days ahead using the methods of extending the transmission parameter *β* described in Section 2.3. The reports are different from those used in the calibration.

| New York City - United States |  |  |  |  |  |  |
| --- | --- | --- | --- | --- | --- | --- |
|  | 10 days | 15 days | 30 days | 45 days | 60 days | 90 days |
| AM | 6.51 (1.90–14.3) | 9.70 (2.44–24.6) | 22.6 (4.98–49.7) | 33.8 (9.23–76.2) | 44.3 (12.3–112) | 63.4 (22.8–215) |
| LR | 7.74 (2.12–16.8) | 11.5 (2.87–26.5) | 24.5 (6.93–52.3) | 35.3 (9.53–79.8) | 44.1 (14.2–119) | 64.1 (19.2–244) |
| TFS | 15.5 (4.77–35.3) | 21.3 (7.21–48.6) | 32.0 (10.8–73.8) | 41.8 (13.9–97.5) | 51.6 (13.4–145) | 72.0 (19.7–247) |
| TFSWN | 15.4 (4.77–35.3) | 21.2 (7.16–48.6) | 32.0 (10.6–73.8) | 41.8 (13.8–97.3) | 51.6 (13.5–144) | 72.0 (19.8–247) |
| MR | 4.84 (1.39–11.2) | 8.23 (2.32–19.5) | 23.2 (6.47–52.0) | 39.4 (13.7–92.4) | 51.0 (20.6–167) | 74.0 (33.1–523) |
| Chicago - United States |  |  |  |  |  |  |
| AM | 7.99 (2.24–18.09) | 12.34 (3.13–27.6) | 28.1 (7.62–67.8) | 49.7 (17.5–110) | 65.9 (24.7–210) | 79.6 (28.2–588) |
| LR | 10.13 (2.79–20.53) | 14.92 (4.52–31.0) | 33.5 (10.3–73.7) | 53.8 (22.2–129) | 69.4 (24.7–242) | 82.1 (28.7–620) |
| TFS | 22.58 (6.45–46.91) | 30.57 (9.78–66.9) | 49.1 (17.7–140) | 68.0 (25.5–269) | 76.4 (31.2–422) | 86.4 (25.3–930) |
| TFSWN | 22.58 (6.54–46.90) | 30.63 (9.79–66.7) | 49.0 (17.8–140) | 68.0 (25.6–268) | 76.3 (31.3–420) | 86.4 (25.1–929) |
| MR | 5.89 (1.86–13.86) | 10.56 (3.14–24.7) | 27.5 (7.67–66.5) | 46.4 (14.6–144) | 61.5 (21.8–310) | 81.8 (37.4–1068) |
| State of Sao Paulo - Brazil |  |  |  |  |  |  |
| AM | 6.76 (1.73–21.9) | 14.6 (3.94–48.6) | 27.3 (7.12–74.7) | 38.4 (10.6–92.8) | 48.0 (14.8–136) | 60.4 (22.8–205) |
| LR | 7.15 (1.80–20.7) | 11.9 (3.36–38.8) | 22.2 (5.79–67.0) | 28.7 (8.90–84.3) | 36.4 (11.3–104) | 50.4 (14.7–206) |
| TFS | 10.2 (2.02–31.2) | 13.0 (2.45–36.8) | 19.0 (5.04–49.9) | 26.1 (7.45–62.6) | 33.2 (8.71–82.3) | 45.7 (13.2–131) |
| TFSWN | 10.3 (2.00–30.7) | 13.0 (2.57–35.5) | 21.1 (5.67–64.8) | 30.4 (8.48–80.9) | 35.9 (11.2–89.9) | 47.5 (14.3–138) |
| MR | 4.98 (1.01–13.7) | 15.6 (4.15–54.2) | 32.9 (9.58–90.0) | 48.0 (16.4–139) | 62.2 (20.9–303) | 74.8 (28.4–748) |
| British Columbia - Canada |  |  |  |  |  |  |
| AM | 7.52 (2.36–20.4) | 12.7 (3.66–32.2) | 27.7 (6.19–68.9) | 43.4 (11.9–117) | 61.2 (16.9–219) | 84.4 (31.8–606) |
| LR | 8.97 (2.74–24.4) | 14.9 (4.61–37.4) | 30.8 (6.66–75.6) | 47.3 (14.1–131) | 64.7 (15.4–248) | 88.3 (28.8–639) |
| TFS | 19.2 (5.16–42.8) | 26.8 (6.87–60.8) | 45.0 (12.2–89.5) | 61.7 (14.7–185) | 73.8 (17.5–311) | 91.2 (30.9–768) |
| TFSWN | 19.2 (5.23–42.8) | 26.9 (6.84–60.8) | 45.0 (12.1–89.5) | 61.7 (14.7–184) | 73.5 (17.4–311) | 91.2 (30.9–769) |
| MR | 5.11 (1.42–14.1) | 9.41 (2.29–27.9) | 22.8 (5.22–67.2) | 36.1 (10.6–136) | 55.2 (15.3–286) | 77.9 (27.9–1479) |

**Figure 1:**
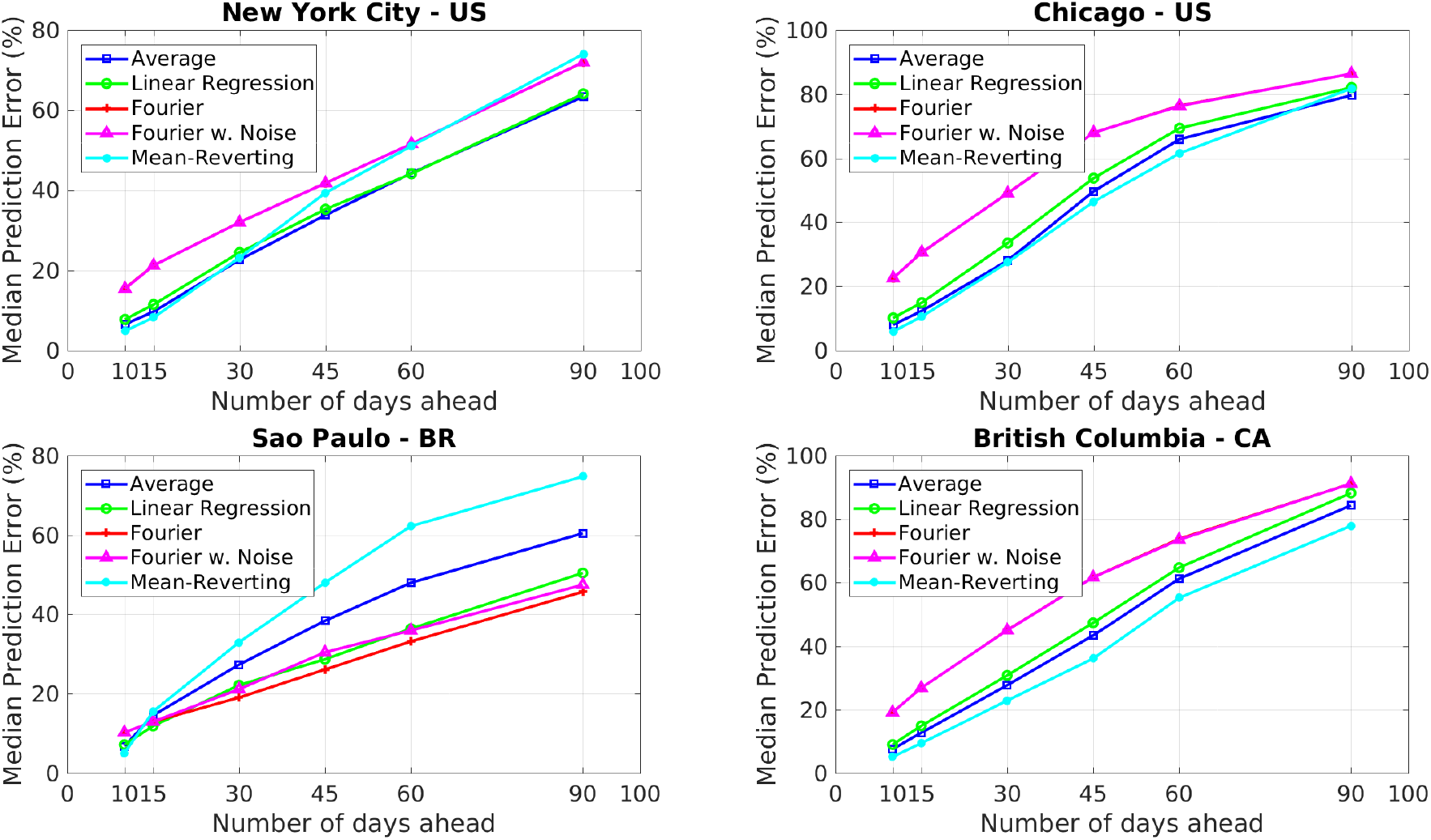
Median values of the percentage error in the out-of-sample predicted accumulated numbers of infections. Predictions are based on calibrated data and evaluated from 10 to 90 days ahead using the methods of extending the transmission parameter *β* described in Section 2.3. The reports are different from those used in the calibration.

To evaluate the errors of the stochastic techniques, namely TFSWN and MR, we used the median values of 5000 samples of predicted infections.

As Figure 1 and Table 1 show, all the methods presented similar error values for predicting 10 to 15 days ahead. Major differences started to appear from 30 to 90 days ahead predictions. Predictions made with the AM presented small median error values with moderate 70% CI even for prediction with larger time intervals. For NYC, Chicago, and BC, it presented one of the smallest median error values for all prediction time intervals. For SP, this technique also presented accurate results, but for longer prediction time intervals, other techniques performed considerably better. Although it is the simplest technique considered in this work, its performance is remarkably accurate.

The LR presented results similar to the AM technique. However, its errors were slightly larger than those for the AM using the datasets from NYC, Chicago, and BC. For SP, it presented a better performance than the AM. It is worth mentioning that such technique is also simple to implement and a nice alternative to the AM.

The TFS and TFSWN presented similar results since their graphs matched for almost all places. They both performed better for SP, but all the other techniques outperformed both. The main reason for such results must be that truncated Fourier series fits the data quite well, which is useful for adherence to data, but may not be the case for model predictions. In other words, it seems that these techniques tend to overfit the data. However, these techniques seems to perform better for longer periods, since, in some sense, they capture better the log-term behavior of the transmission parameter.

For Chicago and BC, the MR outperformed all the other techniques. For NYC it presented one of the best resutls for predictions until 30 days. For SP, it outperformed the other techniques only for predictions until 10 days. In general, for longer periods, like 60 to 90 days, the predictions 70% CIs were wider than those obtained with other techniques. Although, these results may indicate that such technique can be less accurate for larger prediction times, it was effective in predicting the magnitude of new waves of infections for NYC.

In the second example, we focus on the forecast of large outbreaks that happened in New York City during 2020 and 2021, including those caused by the emergence of the delta (third wave) and omicron (fourth wave) variants of the SARS-CoV-2. The four periods considered in this example are from 05-Apr to 17-May-20, from 18-Oct to 29-Nov-20, from 10-Jul to 21-Aug-21, and from 01-Nov to 13-Dec-21, all them with roughly 45 days. In all cases, the calibration uses data until the day before the period of prediction, namely, 04-Apr-20, 17-Oct-20, 09-Jul-21, and 09-Nov-21. Predictions are compared with the accumulated number of reported infections and confidence intervals are generated with bootstrapping [32]. For MR and TFSWN, we use median values of infections. The comparison between reported and predicted accumulated infections during the periods of interest can be found in Figure 2. Each column of panels refers to one outbreak.

**Figure 2:**
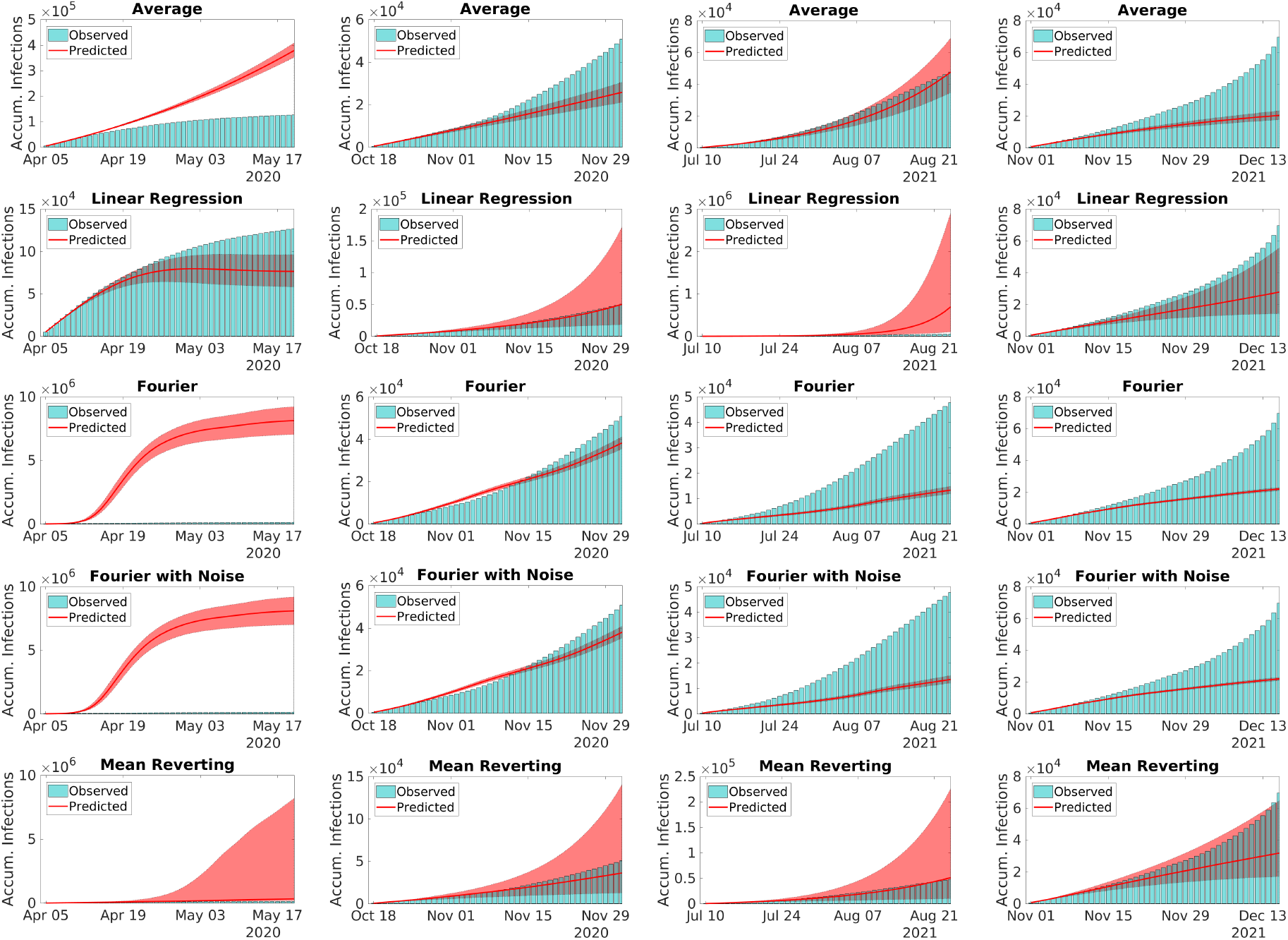
Comparison between reports and model predictions of accumulated COVID-19 infections during 45 consecutive days. Predictions are made just before four outbreaks of COVID-19 in NYC, including those triggered by the Delta and Omicron variants. Bars are the reports, solid lines and the filled areas are the median values and the 90% CI of model predictions, respectively. Predictions are evaluated using the methods for extending the transmission parameter *β* described in Section 2.3.

In the first wave of infections, the LR and MR presented the best forecast performance with median values close to the observed number of infections. The LR started to underpredict the magnitude of the wave after10 to 15 days of forecast, whereas the MR presented a wide 90%CI containing the reports. The AMpresented accurate predictions until 10 to 15 days ahead. After this period, the method over-predicted considerably the reported infections. The TFS and TFSWN forecast poorly the first outbreak probably due to its tendency for overfitting. It is worth noticing that, during the first wave, data was poorer.

In the other three outbreaks, all the models presented similar results. The AM forecast well the magnitude of the three waves, despite of its 90% CI, for the second and fourth waves, did not contained the reported values for periods longer than 20 to 30 days. The LR presented more accurate predictions than the AM. It only overpredicted the the third wave. The TFS and TFSWN predicted well the magnitude of the second wave but underpredicted the other two. The MR predicted accurately all the three waves, with its median values matching the reports of the third wave. The 90% CI was not too wide and contained the reports.

Overall, all the techniques performed well for shorter periods, i.e., from 10 to 15 days. For periods until 45 days, the AM, LR, and MR seem to perform considerably better than the TFS and TFSWN. However, for larger periods of forecast, predictions become less reliable and can be only considered qualitatively.

Although the results of the AM, the LR, and the MR are comparable, the MR has the advantage of incorporating the randomness of the transmission, offering a series of possible future scenarios. Based on these examples we suggest that, for shorter periods of forecast, i.e., until 45 days, the AM and MR are reliable choices. For longer periods, their results must be compared with those given by the TFS or the TFSWN. The advantage of the TFSWN over the TFS, is the incorporation of uncertainty, that make it easy to provide predictions naturally inside confidence intervals.

To illustrate that the results above can be extended to a larger class of SEIR-type models, we repeat the first experiment with NYC data and the model proposed by Albani et al. [3]. The version of the model used here accounts for different levels of disease severity and their dependency on age-range. Figure 3 shows the evolution of the median values of the normalized forecast errors. The same values with their 70% CI can be found in Table 2.

**Table 2:** Median values and their corresponding 70% confidence intervals of the percentage error in the predicted accumulated numbers of infections. Predictions are based on calibrated data and evaluated from 10 to 90 days ahead using the methods of extending the transmission parameter *β* described in Section 2.3.

| New York City - United States |  |  |  |  |  |  |
| --- | --- | --- | --- | --- | --- | --- |
|  | 10 days | 15 days | 30 days | 45 days | 60 days | 90 days |
| AM | 4.60 (1.89–8.73) | 8.09 (4.27–18.6) | 19.0 (7.69–56.0) | 29.8 (11.0–70.5) | 37.7 (11.8–124) | 58.7 (11.2–344) |
| LR | 5.78 (2.34–9.35) | 10.6 (4.63–16.5) | 24.1 (11.7–43.1) | 34.9 (11.8–70.0) | 42.8 (10.7–125) | 63.4 (21.6–211) |
| TFS | 8.29 (3.30–15.0) | 12.7 (5.16–22.1) | 27.5 (4.50–53.7) | 35.7 (11.0–75.0) | 44.8 (23.7–114) | 60.2 (32.5–191) |
| TFSWN | 8.43 (3.39–15.0) | 13.1 (5.18–22.3) | 27.5 (4.69–53.9) | 35.9 (10.5–75.0) | 44.2 (23.3–114) | 60.3 (34.2–191) |
| MR | 6.91 (1.76–14.2) | 11.1 (5.14–23.1) | 29.8 (5.07–51.5) | 47.7 (17.1–115) | 51.6 (20.6–448) | 86.2 (36.1–1532) |

**Figure 3:**
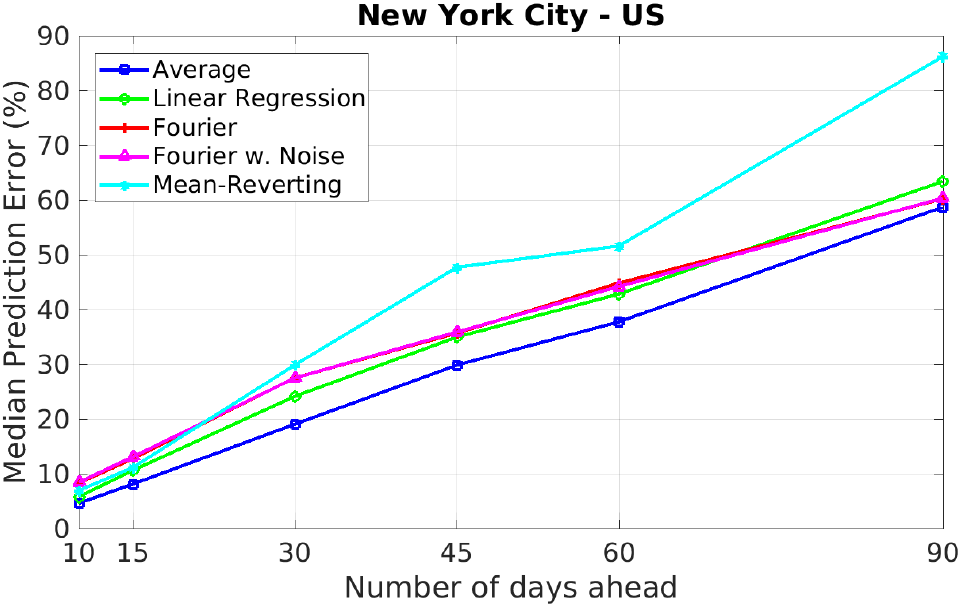
Median values of the percentage error in the predicted accumulated numbers of infections. Predictions are based on calibrated data and evaluated from 10 to 90 days ahead using the methods of extending the transmission parameter *β* described in Section 2.3. The epidemiological model in Albani et al. [3] was used.

As Figure 3 and Table 2 show, by including more characteristics of the disease in the SEIR-type model, the forecast overall accuracy did not improve. We used all the forecast techniques from Section 2.3. All of them presented similar forecast errors. Such pattern may be linked to the increase of the uncertainties in model calibration, since more sophisticated models have larger sets of unknowns for the same experimental datasets, since NYC does not have daily reports of COVID-19 cases, hospitalizations, and deaths classified by age-range.

## 4 Discussion

Although the model used to test different predictions techniques may seem to be simple, by considering time varying transmission parameter and death rate, it incorporates naturally stochastic fluctuations in the pathogen transmission, like the introduction of new variants, changes in the disease virulence and mortality. In other words, providing an accurate nowcast. More sophisticated models are harder to calibrate and to forecast scenarios within longer periods, it is necessary to predict the evolution of other quantities, like mobility information or the disease severity dependency on age. It is also difficult to estimate the causality relations between transmission and mobility [33, 34]. Moreover, as Figure 3 shows, the performance of more sophisticated models may not be considerably better. Thus, if the main interest is in scenarios regarding the general population, it is better to use simpler, but robust and accurate, models.

Using the same dataset, predictions with different techniques presented considerably different performances in forecast. For shorter periods, the AM, the LR, and the MR outperformed the TFS and the TFSWN. It is also interesting that the MR predicted accurately the four waves of infections in NYC, as illustrated by Figure 2. In all cases, the model prediction 90% CI contained the reported infections. This indicates that the MR incorporates better the uncertain nature of the virus transmission and shows that transmission parameter *β* must be modeled as a stochastic process.

Reliability of model forecast decreases considerably with respect to the length of the prediction time interval, as Tables 1–2 show. This is due to unpredictable regime changes in the transmission dynamics, like the emergence of new strains of the pathogen, vaccination, or yet effectiveness of non-pharmaceutical measures like lockdowns. Thus, for periods longer than 45 days, predictions must be considered only qualitatively.

Finally, the present study has some limitations since we considered only the class of SEIR-type models. There are a number of other ways to model the dynamics of infectious diseases [2], but accounting for them is beyond the scope of this article that is intended to enlighten the stochastic nature of transmission. Moreover, we did not perform a rigorous statistical analysis of the transmission parameter and other rates defining the SEIR-type model in Eqs. (2)–(5) since the calibration techniques considered in this work are deterministic. The statistical analysis of SEIR-type models and their parameters will be subject of a future work.

## Data Availability

All data produced in the present study are available upon reasonable request to the authors. All the codes used to produce the data are vailable online at GitHub.

https://github.com/viniciusalbani/NowcastForecastCovid

## Data Availability

The datasets used in this study are publicly available [35–38].

## Code Availability

The codes in MATLAB (The MathWorks, Inc., Natick, USA) used in this work can be found in https://github.com/viniciusalbani/NowcastForecastCovid.

## Acknowledgments

VA acknowledges the financial support from Fundação Butantan and Fundação de Amparo à Pesquisa e Inovação do Estado de Santa Catarina through the grants 01/2020 and 00002847/2021, respectively. RA acknowledges the financial support from Fundação Carlos Chagas Filho de Amparo à Pesquisa do Estado do Rio de Janeiro (FAPERJ) through the grants E-26/202.932/2019 and E-26/202.933/2019. EM acknowledges the financial support from Conselho Nacional de Desenvolvimento Científico e Tecnològico (CNPq) and Fundação Butantan through the Grants 305544/2011-0 and 01/2020, respectively. JZ acknowledges the financial support from Khalifa University, CNPq, and Fundação Carlos Chagas Filho de Amparo à Pesquisa do Estado do Rio de Janeiro through the Grants FSU-2020-09, 307873/2013-7, and E-26/202.927/2017, respectively.

## Author contributions statement

All the authors contributed equally to this work.

